# An exploratory analysis of circulating cytokine profiles, pain-associated transcripts and autoantibody responses associated with treatment response in knee osteoarthritis

**DOI:** 10.64898/2026.10.25.26365740

**Authors:** Susan Franks, Afroditi Kouraki, Tony Kelly, Richard Berwick, Harvey Neiland, Andreas Goebel, Ana M Valdes

## Abstract

**Background:** Pain response to treatment in knee osteoarthritis (OA) varies substantially between individuals, but the biological factors underlying this heterogeneity remain poorly understood. We investigated whether circulating protein, transcriptional and autoantibody features were associated with variation in pain response to lifestyle interventions in people with knee OA.

**Methods:** We performed an exploratory multi-modal analysis using participants from the INSPIRE randomised controlled trial and, for proteomic analyses, an independent WebEx cohort. Circulating inflammatory proteins were measured using Olink proximity extension assay at baseline and 6 weeks. Associations between baseline protein concentrations, longitudinal protein changes and pain response were assessed using multivariable regression, with multivariate protein patterns examined using supervised and sparse partial least squares approaches. Whole-blood transcriptomic data from INSPIRE were analysed using limma, restricted a priori to pain-associated genes represented in the Human Pain Genetics Database (HPGDB), with a responder-by-visit interaction used to assess differential longitudinal expression. Finally, exploratory IgG binding to murine satellite glial cells (SGCs) was assessed in selected INSPIRE participants with contrasting clinical responses to exercise.

**Results:** No circulating protein or transcriptomic association remained significant after correction for multiple testing. Several nominal associations were observed in the proteomic analyses, including baseline IL-10RA and CST5 in INSPIRE and OSM and TNFSF14 in WebEx. Longitudinal changes in IL-17A and TSLP were nominally associated with improvement in pain in INSPIRE, with a similar nominal association between IL-17A and pain improvement observed in WebEx. In the transcriptomic analysis, MMP9 showed the strongest nominal association with differential longitudinal expression according to pain response under both the ΔNRS ≤ −1 (log2 fold change = 0.91, p = 0.008) and ΔNRS ≤ −2 (log2 fold change = 1.02, p = 0.004) definitions; neither association remained significant following FDR correction. Exploratory SGC-binding analysis identified markedly increased baseline IgG binding in one participant with worsening pain, but this pattern was not consistently observed among other participants with poor clinical responses. Paired analyses provided no evidence of a consistent increase in SGC-binding IgG following exercise.

**Conclusions:** This exploratory multi-modal analysis found no robust molecular signature of the pain response that survived correction for multiple testing. Nominal associations involving several circulating proteins, including IL-10RA, IL-17A and MMP9 provide potential leads for future investigation, but these findings should be interpreted cautiously given the multiple comparisons and limited sample sizes. Exploratory autoantibody analyses likewise provided no evidence of a consistent SGC-binding signature. Larger, independently powered studies are required to determine whether these candidate molecular features are reproducibly associated with treatment-related variation in pain response.

## Introduction

Pain is a major contributor to disability and reduced quality of life in people with osteoarthritis (OA) [1]. Although interventions such as exercise and physiotherapy can provide clinically meaningful improvements in pain [2,3], treatment response varies substantially between individuals [4]. In our previously published randomised controlled trial, INSPIRE, both inulin supplementation and physiotherapy-supported exercise (PSE) resulted in clinically meaningful reductions in knee pain after 6 weeks, while inulin supplementation was additionally associated with improvements in pain sensitisation [3]. Exploratory analysis of the gut microbiome from the same trial suggested that baseline microbial community composition may be associated with subsequent pain improvement [5]. However, the biological factors underlying variation in pain response remain poorly understood.

Inflammatory and immune signalling pathways have been implicated in the development and maintenance of OA-related pain [6]. Cytokines and other immune-related proteins can modulate peripheral nociceptor sensitivity, neuroimmune interactions and tissue inflammation, and alterations in these pathways may accompany changes in pain. Circulating proteins may therefore provide insight into biological processes associated with treatment response and potentially identify molecular profiles that distinguish individuals who experience clinically meaningful pain improvement from those who do not. However, the relationship between circulating inflammatory protein profiles and clinical pain response remains unclear.

Beyond circulating proteins, genetic and transcriptomic evidence suggests that individual variation in pain may reflect differences in nociceptive and neuroimmune pathways. The Human Pain Genetics Database (HPGDB) integrates evidence from human genetic studies and identifies a broad set of genes associated with pain phenotypes, including relating to pathways involved in nociception, analgesia and inflammation [7]. Functional genomic studies have further highlighted the value of transcriptomic approaches for linking genetic susceptibility to molecular mechanisms in OA and related pain phenotypes [8]. We therefore used a predefined set of pain-associated genes from the HPGDB to examine whether longitudinal transcriptional changes were associated with clinical pain response. In parallel, emerging evidence implicates adaptive immune mechanisms, with circulating autoantibodies, B-cell responses and other features of self-reactivity reported in subsets of people with OA [9]. This raises the possibility that antibodies directed towards neural or neuroimmune targets may contribute to individual differences in pain phenotypes. In particular, patient-derived IgG has been shown to bind satellite glial cells and sensory neurons and to induce pain-related hypersensitivity in experimental models of fibromyalgia [10]. We therefore explored SGC-binding IgG as a potential neuroimmune correlate of poor or paradoxical pain responses to exercise.

In the present study, we investigated biological correlates of inter-individual variation in pain response to lifestyle interventions in people with knee OA. We first characterised circulating protein profiles using Olink proximity extension assay in participants from the INSPIRE randomised controlled trial, examining both baseline protein concentrations and longitudinal changes in relation to pain response. Findings were subsequently examined in the independent WebEx cohort where appropriate. To investigate whether transcriptional differences in established pain-related genes accompanied variation in clinical response, we additionally analysed whole-blood transcriptomic data using a predefined set of pain-associated genes from the HPGDB. Finally, to explore whether adaptive immune mechanisms might contribute to poor or paradoxical responses to exercise, we performed an exploratory satellite glial cell-binding assay using purified IgG from selected INSPIRE participants with contrasting clinical responses. Together, these complementary analyses were used to explore circulating protein, transcriptional and autoantibody features associated with heterogeneity in response to exercise-based and nutritional interventions in knee OA. Pain responses were categorised using two predefined thresholds based on reductions in Numerical Rating Scale (NRS) pain scores of ≥1 and ≥2 points, allowing assessment of whether associations were consistent across definitions of clinically meaningful improvement.

## Methods

### Study population

Data were derived from two randomised controlled trials of interventions for knee osteoarthritis (OA), INSPIRE and WebEx. The primary analysis used data from the INSPIRE 2 × 2 factorial randomised controlled trial (ClinicalTrials.gov: NCT05670314), which investigated whether prebiotic nutritional supplementation, alone or in combination with physiotherapy-supported exercise (PSE), could improve pain outcomes through distinct or complementary gut microbiome mechanisms. Participant recruitment, eligibility criteria, intervention details and outcome assessment have been described previously (3). Briefly, participants with knee OA were randomised to receive inulin supplementation (20 g/day), web-based PSE, both interventions, or placebo maltodextrin (10 g/day) for 6 weeks. The primary outcome was knee pain severity, measured using the Numerical Rating Scale (NRS), with participants rating average pain over the preceding week from 0 (no pain) to 10 (worst imaginable pain). Clinical assessments and biological samples were collected at baseline and 6 weeks post-intervention. The INSPIRE trial was approved by the University of Nottingham Faculty of Medicine & Health Sciences Research Ethics Committee (Ref: 473-0322), and all participants provided written informed consent. The present analysis included 89 participants with both clinical pain outcomes and Olink proteomic measurements.

An independent cohort from the Web-Based Exercises for Treating Knee Osteoarthritis (WebEx) randomised controlled trial (ClinicalTrials.gov: NCT03545048) was additionally analysed to assess the consistency of the observed protein–pain associations. WebEx evaluated the effectiveness of internet-based exercise in individuals with knee OA, with NRS pain scores assessed at baseline and post-intervention [11]. Olink proteomic measurements were available for 51 participants, who were included in the present analysis.

### Cytokine profiling

Fasting blood samples were collected at baseline and follow-up visits by trained staff using 6 mL Gold Vacutainer serum separation tubes. After resting for 30 min, the samples were centrifuged to obtain serum aliquots and stored in −80°C freezers following local standard operating procedures. Inflammatory protein measurements were performed using a proximity extension assay method [12] with the Olink inflammatory panel [13].

### Responder definitions

Two binary pain improvement outcomes were defined for INSPIRE using changes in NRS pain scores from baseline. Participants were classified as responders if their pain scores decreased by 1 point or more (ΔNRS ≤ -1, n = 56 responders vs 33 non-responders) or 2 points or more (ΔNRS ≤ -2, n = 35 responders vs 54 non-responders), and non-responders otherwise. These thresholds were respectively chosen to represent minimal and moderate clinically meaningful improvements in pain intensity. For WebEx, responders were defined by ΔNRS ≤ -1 (n = 36 responders vs 15 non-responders).

### Baseline and longitudinal linear models

Binomial generalised linear models were fitted to assess the associations between binary outcomes (0/1) of pain response and (i) baseline protein concentrations and (ii) changes in protein concentrations. Models were adjusted for the covariates (age, sex, body mass index (BMI) and treatment arm).

Associations between changes in protein concentrations and continuous changes in pain were assessed using linear regression models adjusted for covariates. Given the large number of proteins assessed, p-values were adjusted for multiple testing using the Benjamini–Hochberg false discovery rate (FDR) correction and a FDR-adjusted p-value of less than 0.05 was considered statistically significant.

### Multivariate analysis

Supervised partial least squares discriminant analysis (sPLS-DA) was performed using the mixOmics package in R to identify whether combinations of baseline cytokines differentiated between pain responders and non-responders. Cytokine concentrations were centred and scaled prior to analysis. Two components were fitted, with 10 cytokines retained per component. Model performance was assessed using five-fold M-fold cross-validation. Variable-selection stability was evaluated using 500 repeated 80% subsamples, with selection frequency used to identify consistently selected cytokines.

Multivariate associations between longitudinal changes in cytokine concentrations and changes in pain were assessed using sparse partial least squares (sPLS) regression. Change in pain was adjusted for covariates and cytokine changes were centred and scaled prior to analysis. Two components were fitted, with 10 cytokines retained per component. Variable-selection stability was assessed using 500 repeated 80% subsamples, with selection frequency used to identify consistently selected cytokines.

### Transcriptomic analysis of pain-associated genes

Whole-blood transcriptomic data generated from INSPIRE participants were analysed to investigate whether transcriptional differences in genes previously implicated in pain biology were associated with clinical pain response. Transcriptomic RNA sequencing data were processed and cleaned by GeneWiz. Sequencing reads were trimmed with Trimmomatic v0.36 and aligned to the Homo sapiens GRCh38 reference genome (ENSEMBL) using STAR v2.5.2b, which incorporates splice junctions for accurate alignment. Unique exon-aligned reads were then quantified using featureCounts (Subread v1.5.2) with the gene_id annotation, applying strand-specific counting when appropriate.

We performed analysis restricted a priori to genes represented in the HPGDB, thereby focusing the analysis on genes with previously reported genetic or biological links to pain phenotypes. Raw gene-level count data were analysed using the limma framework in R and were filtered to remove lowly expressed genes and transformed using the voom method to estimate the mean–variance relationship and generate precision-weighted expression values. Differential longitudinal expression according to pain-response status was assessed using a linear model incorporating pain-response status, visit, their interaction, age, sex and BMI. To account for repeated measurements from the same participant, within-participant correlation was estimated using the *duplicateCorrelation* approach and incorporated into the linear model. The primary contrast of interest was the interaction between pain-response status and visit, representing the difference in change in gene expression from baseline to follow-up between pain responders and non-responders. Analyses were performed separately for the two predefined pain-response thresholds (ΔNRS ≤ −1 and ΔNRS ≤ − 2). P-values were adjusted for multiple testing using the Benjamini–Hochberg procedure, with an FDR-adjusted P-value < 0.05 considered statistically significant.

### Exploratory assessment of SGC-binding autoantibodies

To investigate whether autoantibody-mediated mechanisms might contribute to heterogeneity in pain response following exercise, IgG binding to satellite glial cells (SGCs) was assessed in a subset of INSPIRE participants. The assay was adapted from the previously described method by Berwick et al. [10], which demonstrated binding of patient-derived IgG to SGCs and sensory neurons in the dorsal root ganglia.

Serum samples were obtained at baseline and, where available, following the 6-week intervention. IgG was purified from serum and used for immunocytochemical assessment of binding to murine SGCs, following the previously published approach. IgG from a patient with fibromyalgia and established SGC binding was included as a positive control, together with healthy control samples. SGC binding was quantified from fluorescence images using pixel-intensity measures, consistent with the quantitative approach described previously by Berwick et al. [10].

Participants were selected for exploratory analysis based on contrasting clinical responses to the exercise intervention, including participants with marked worsening or absence of improvement in pain and/or pressure pain sensitivity, as well as a participant demonstrating clear clinical improvement. Because the number of participants was small and the assay was performed as an exploratory mechanistic investigation, results were interpreted descriptively rather than as a powered comparison between clinical response groups.

## Results

### Baseline protein profiles and pain response

Across both cohorts, no individual baseline protein association remained significant following correction for multiple testing. In INSPIRE, binomial generalised linear models identified nominal associations between baseline protein concentrations and pain responder status. Higher baseline IL-10RA concentrations were associated with non-response for both definitions of pain response (p = 0.013 and p = 0.028, respectively), while higher CST5 concentrations were associated with response using the more stringent ΔNRS ≤ −2 definition (**Table 1**).

**Table 1:** Association of pain response status (ΔNRS ≤ −1 and ΔNRS ≤ −2) with baseline cytokine levels adjusted for covariates for INSPIRE cohort.

| Pain response | term | estimate | std.error | statistic | p.value | conf.low | conf.high | FDR |
| --- | --- | --- | --- | --- | --- | --- | --- | --- |
| $\Delta\text{NRS} \leq -1$ | IL-10RA | -0.48183 | 0.193574 | -2.48911 | 0.012807 | -0.92028 | -0.15675 | 0.934005 |
| $\Delta\text{NRS} \leq -2$ | IL-10RA | -0.28528 | 0.129892 | -2.19632 | 0.028069 | -0.57356 | -0.05432 | 0.942057 |
|  | CST5 | 0.600385 | 0.300623 | 1.997133 | 0.045811 | 0.031336 | 1.222956 | 0.942057 |

Consistent with these findings, in the sPLS-DA analysis, Component 1 primarily differentiated pain non-responders from responders, with non-responders characterised by higher baseline levels of several inflammatory and immune-related proteins. For the ΔNRS ≤ −1 responder definition (**Table 2**), IL-10RA and CXCL11 were the principal contributors to Component 1, showing high stability of selection across repeated subsampling (100% and 92%, respectively). For the ΔNRS ≤ −2 definition (**Table 2**), IL-10RA and IFN-γ were higher in non-responders, with selection frequencies of 95% and 92%, respectively. IL-17A and IL-1α were also selected, although with lower stability (73% and 64%, respectively). These selection frequencies indicate stability of variable selection rather than statistical significance, and the multivariate findings did not provide evidence of an FDR-significant protein signature.

**Table 2:** Stability of baseline protein selection in sPLS-DA for the ΔNRS ≤ −1 and ΔNRS ≤ −2 pain responder definition denoted by selection frequencies for proteins selected on Component 1 across 500 repeated 80% subsamples for INSPIRE cohort.

| Pain response | Cytokine | Selection | Mean Loading | Median Loading | SD | Lower95 | Upper95 |
| --- | --- | --- | --- | --- | --- | --- | --- |
| **$\Delta\text{NRS} \leq -1$** | IL.10RA | 1 | -0.395 | -0.624 | 0.543 | -0.881 | 0.799 |
|  | CXCL11 | 0.916 | -0.208 | -0.274 | 0.313 | -0.664 | 0.499 |
| **$\Delta\text{NRS} \leq -2$** | IL.10RA | 0.952 | 0.409 | 0.491 | 0.367 | -0.669 | 0.868 |
|  | IFN.gamma | 0.828 | 0.246 | 0.254 | 0.268 | -0.415 | 0.683 |
|  | IL.17A | 0.734 | 0.222 | 0.213 | 0.252 | -0.317 | 0.675 |
|  | IL.1.alpha | 0.636 | 0.152 | 0.068 | 0.212 | -0.134 | 0.673 |

To assess the consistency of these findings in an independent cohort, the same analyses were performed in WebEx. In WebEx, higher baseline concentrations of OSM and TNFSF14 showed nominal associations with non-responders (p = 0.006, p = 0.009, respectively (**Table 3**)). However, neither of these associations remained significant after correction for multiple testing.

**Table 3:** Association of pain response status with baseline cytokine levels adjusted for covariates for WebEx cohort.

| term | estimate | std.error | statistic | p.value | conf.low | conf.high | FDR |
| --- | --- | --- | --- | --- | --- | --- | --- |
| OSM | -1.25966 | 0.456065 | -2.76202 | 0.005744 | -2.25717 | -0.43337 | 0.39394 |
| TNFSF14 | -1.60704 | 0.611276 | -2.62899 | 0.008564 | -2.93443 | -0.49842 | 0.39394 |

The sPLS-DA analysis identified a multivariate baseline protein profile differentiating pain non-responders from responders. Component 1 was characterised by higher levels of several inflammatory and immune-related proteins in non-responders, with OSM, TNFSF14 and CCL20 showing the greatest stability of selection across repeated subsampling (100%, 100% and 92%, respectively (**Table 4**)). As in INSPIRE, these multivariate findings should be interpreted as exploratory patterns of variable selection rather than evidence of statistically significant protein associations.

**Table 4:** Stability of baseline protein selection in sPLS-DA for pain response denoted by selection frequencies for proteins selected on Component 1 across 500 repeated 80% subsamples for WebEx cohort.

| Cytokine | Selection | MeanLoading | MedianLoading | SD | Lower95 | Upper95 |
| --- | --- | --- | --- | --- | --- | --- |
| OSM | 0.996 | -0.394 | -0.555 | 0.412 | -0.743 | 0.635 |
| TNFSF14 | 0.996 | -0.335 | -0.477 | 0.403 | -0.738 | 0.689 |
| CCL20 | 0.922 | -0.196 | -0.253 | 0.285 | -0.630 | 0.518 |

### Longitudinal changes in protein concentrations and pain response

No longitudinal protein association remained significant following FDR correction in either cohort. In INSPIRE, linear regression identified changes in IL-17A and TSLP to be nominally associated with changes in pain (**Table 5**). Specifically, increases in both cytokines were associated with greater improvement in pain.

**Table 5:** Association of changes in pain with changes in protein concentrations adjusted for covariates for INSPIRE cohort.

| term | estimate | std.error | statistic | p.value | conf.low | conf.high | FDR |
| --- | --- | --- | --- | --- | --- | --- | --- |
| IL.17A | -0.60249 | 0.266575 | -2.26012 | 0.026498 | -1.13289 | -0.07209 | 0.999172 |
| TSLP | -0.10007 | 0.048201 | -2.07603 | 0.041063 | -0.19597 | -0.00416 | 0.999172 |

Examining whether longitudinal changes in cytokine concentrations were associated with pain responder status indicated that for the ΔNRS ≤ −1 definition (**Table 6**), greater increases in ARTN, a protein implicated in nociceptive signalling, were associated with responder status. Using the more stringent ΔNRS ≤ −2 definition (**Table 6**), increases in IL-22RA1, the receptor for IL-22, were associated with non-response. Neither of these associations remained significant after FDR correction.

**Table 6:** Association of pain responder status (ΔNRS ≤ −1 and ΔNRS ≤ −2) with changes in protein concentrations adjusted for covariates for INSPIRE cohort.

| Pain response | term | estimate | std.error | statistic | p.value | conf.low | conf.high | FDR |
| --- | --- | --- | --- | --- | --- | --- | --- | --- |
| $\Delta\text{NRS} \leq -1$ | ARTN | 1.29172 | 0.639102 | 2.021149 | 0.043264 | 0.12294 | 2.664866 | 0.917466 |
| $\Delta\text{NRS} \leq -2$ | IL.22.RA1 | -0.71908 | 0.333853 | -2.15388 | 0.031249 | -1.431 | -0.10819 | 0.635506 |

Component 1 in the sPLS regression was characterised predominantly by lower changes in IL-17A, TSLP, TNF, IL-6 and ARTN, with higher Component 1 scores associated with less improvement in pain. Among these proteins, IL-17A and TSLP showed the greatest stability of selection, being selected in 68% and 58% of repeated subsamples, respectively (**Table 7**). TNF, IL-6 and ARTN were selected in fewer than 45% of repeated subsamples, suggesting that selection stability across the multivariate model was generally modest.

**Table 7:** Stability of longitudinal protein selection in sPLS regression determined from selection frequencies and loading estimates across 500 repeated 80% subsamples. Protein changes from the INSPIRE cohort were centred and scaled, and change in pain was adjusted for covariates.

| Cytokine | Selection | MeanLoading | MedianLoading | SD | Lower95 | Upper95 |
| --- | --- | --- | --- | --- | --- | --- |
| IL-17A | 0.676 | -0.291 | -0.269 | 0.272 | -0.810 | 0 |
| TSLP | 0.578 | -0.249 | -0.127 | 0.286 | -0.844 | 0 |
| TNF | 0.418 | -0.121 | 0 | 0.184 | -0.592 | 0 |
| IL6 | 0.342 | -0.095 | 0 | 0.171 | -0.573 | 0 |
| ARTN | 0.324 | -0.123 | 0 | 0.225 | -0.753 | 0 |

In WebEx, changes in IL-17A showed a nominal association with changes in pain, with increases associated with greater improvement in pain (**Table 8**). This direction of association was consistent with the finding in INSPIRE, although the association in WebEx also did not remain significant following FDR correction. Changes in TNF-β, IFN-γ, IL-2RB, CD6 and FGF-23 were also nominally associated with pain responder status, with greater increases observed in non-responders (**Table 9**). None of these associations remained significant following FDR correction.

**Table 8:** Association of changes in pain with changes in protein concentrations adjusted for covariates for WebEx cohort.

| term | estimate | std.error | statistic | p.value | conf.low | conf.high | FDR |
| --- | --- | --- | --- | --- | --- | --- | --- |
| IL-17A | -1.32452 | 0.64091 | -2.06663 | 0.044553 | -2.61538 | -0.03366 | 0.449963 |

**Table 9:** Association of pain responder status with changes in protein concentrations adjusted for covariates for WebEx cohort.

| term | estimate | std.error | statistic | p.value | conf.low | conf.high | FDR |
| --- | --- | --- | --- | --- | --- | --- | --- |
| TNFB | -2.24736 | 1.028398 | -2.1853 | 0.028867 | -4.57357 | -0.45608 | 0.372726 |
| IL2RB | -2.65353 | 1.24567 | -2.1302 | 0.033155 | -5.4403 | -0.45964 | 0.372726 |
| FGF.23 | -2.25193 | 1.110115 | -2.02856 | 0.042503 | -4.68839 | -0.23715 | 0.372726 |
| IFN.gamma | -0.92162 | 0.455482 | -2.0234 | 0.043032 | -1.94449 | -0.11822 | 0.372726 |
| CD6 | -1.73321 | 0.877676 | -1.97477 | 0.048294 | -3.67547 | -0.25059 | 0.372726 |

In the longitudinal sPLS regression analysis, Component 1 was characterised predominantly by lower changes in IL-17A, ADA, CCL4 and SCF, with higher Component 1 scores associated with less improvement in pain (**Table 10**). IL-17A showed the greatest stability of selection across repeated subsampling, being selected in 53% of subsamples, although overall selection stability was low.

**Table 10:** Stability of longitudinal protein selection in sPLS regression determined from selection frequencies and loading estimates across 500 repeated 80% subsamples. Protein changes from the WebEx cohort were centred and scaled, and change in pain was adjusted for covariates.

| Cytokine | Selection | MeanLoading | MedianLoading | SD | Lower95 | Upper95 |
| --- | --- | --- | --- | --- | --- | --- |
| IL.17A | 0.526 | -0.241 | -0.046 | 0.290 | -0.829 | 0 |
| ADA | 0.476 | 0.135 | 0 | 0.201 | 0 | 0.691 |
| CCL4 | 0.454 | 0.142 | 0 | 0.200 | 0 | 0.614 |
| SCF | 0.388 | 0.097 | 0 | 0.165 | 0 | 0.523 |

**Table 11.** INSPIRE study patients included in IgG binding analyses. Pain intensity was self-reported on a 0–10 scale, and peripheral sensitisation was assessed via brachioradialis pressure pain threshold (PPT).

| Patient | OA ID | Baseline pain NRS | Post-exercise pain NRS | Change in Brachioradialis PPT (kPa; week 6 – baseline) |
| --- | --- | --- | --- | --- |
| Pt1 | OA 1 | 6 | 9 | -41.2 |
| Pt2 | OA 2 | 7 | 6 | +102.9 |
| Pt3 | OA 3 | 7 | 6 | -24.5 |
| Pt4 | OA 4 | 5 | 9 | -12.8 |
| Pt5 | OA 5 | 8 | 4 | +39.2 |

Brachioradialis pressure pain threshold (PPT) was measured at baseline and week 6. Change was calculated as the week 6 value minus the baseline value (kPa). Positive values indicate an increase in PPT, reflecting reduced pressure pain sensitivity, whereas negative values indicate a decrease in PPT, reflecting increased pressure pain sensitivity.

### Transcriptomic analysis of pain-associated genes

To investigate whether transcriptional differences in established pain-associated genes accompanied variation in clinical response, whole-blood transcriptomic data were analysed using a predefined set of genes from the HPGDB. No genes remained significantly associated with either pain-response definition following correction for multiple testing.

Among the genes examined, MMP9 showed the strongest nominal association with pain response under both responder definitions. For the ΔNRS ≤ −1 definition, MMP9 showed a greater longitudinal increase in the responder group relative to the non-responder group (log2 fold change = 0.91, p = 0.008), while a similar association was observed using the more stringent ΔNRS ≤ −2 definition (log2 fold change = 1.02, p = 0.004). Neither association remained significant following false discovery rate correction. Other genes showed weaker nominal associations under the individual response definitions, but these were not consistent across both thresholds.

### Exploratory assessment of SGC-binding autoantibodies

We next explored whether SGC-binding IgG was associated with variation in response to exercise. As we considered that minor changes in pain intensity may not reflect in this test, for this exploratory analysis we selected patients with full data sets available who had at least a pain intensity of NRS 5 and who showed either dramatic improvement, including with their pressure pain threshold (PPT), dramatic worsening, or relatively little change (defined for the purpose of this test as no or +/-1 NRS point change). All patients fulfilling these criteria were included. Baseline and post-exercise pain scores and PPT values are reported for all selected patients.

PPT was assessed using an algometer, as described previously [3], and was defined as the minimum pressure required to elicit pain. Brachioradialis PPT was measured on the arm contralateral to the painful knee, providing an assessment of pressure pain sensitivity at a site distant from the primary pain location.

Baseline IgG binding to murine SGCs was initially examined in three INSPIRE participants with poor clinical responses to exercise and three healthy controls, with IgG from a patient with fibromyalgia and established SGC binding included as a positive control. Healthy controls demonstrated low levels of IgG binding, whereas one participant (OA1/Pt1) showed markedly increased binding, comparable to the positive control. In contrast, OA2 and OA3 showed binding levels similar to those observed in healthy controls (**Figure 1**).

**Figure 1.**
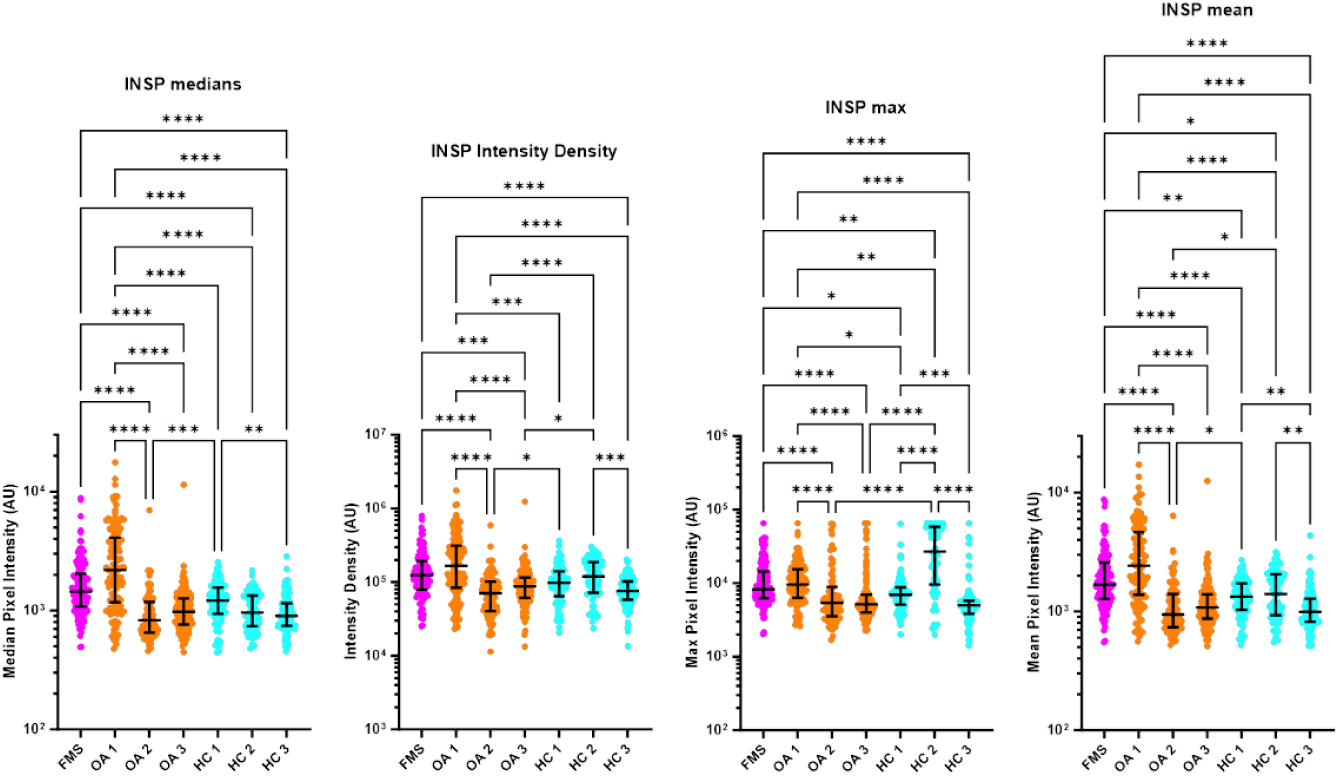
Baseline IgG binding to murine SGCs was assessed for three OA non/minimal-responders (OA 1 = Pt1, OA 2 = Pt2, OA 3 = Pt3), one FMS patient (positive control), and three healthy controls. Median pixel intensity was used to quantify binding. All HCs showed low binding.

We subsequently examined OA4, who experienced a marked increase in pain following exercise. Contrary to the initial hypothesis that baseline SGC-binding autoantibodies might identify participants who subsequently develop increased pain, OA4 demonstrated low baseline IgG binding, lower than the other OA participants tested (**Figure 2**). Thus, elevated baseline SGC binding was not consistently associated with worsening clinical symptoms.

**Figure 2.**
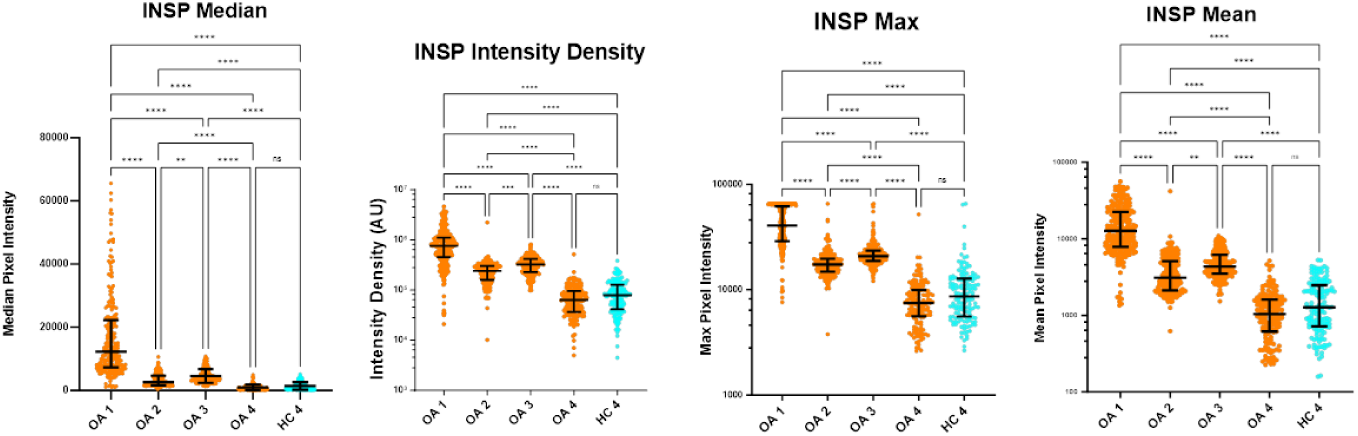
Baseline IgG binding from OA 1 (Pt1), OA 2 (Pt2), OA 3 (Pt3), and OA 4 (Pt4) was assessed using baseline serum. OA 1-3 demonstrated the same relative IgG binding in experiment 1. Although OA 4’s pain worsened from 5 to 9 post-exercise, baseline IgG binding was very low.

To determine whether exercise might instead modulate an adaptive immune response, paired baseline and post-intervention samples were subsequently analysed. Across the participants examined, there was no evidence for any pattern of response in SGC-binding IgG following exercise that would reflect recorded changes in pain intensity. Although some maximum and mean pixel-intensity measures differed between visits for OA1 (**Figure 3**), the median pixel intensity did not differ significantly, and similar patterns were observed in subsequent experiments involving OA3 and OA4 (**Figure 4**). Overall, these exploratory analyses provided no evidence of a consistent baseline SGC-binding IgG phenotype associated with poor clinical response, nor of a consistent change in SGC-binding IgG following exercise.

**Figure 3.**
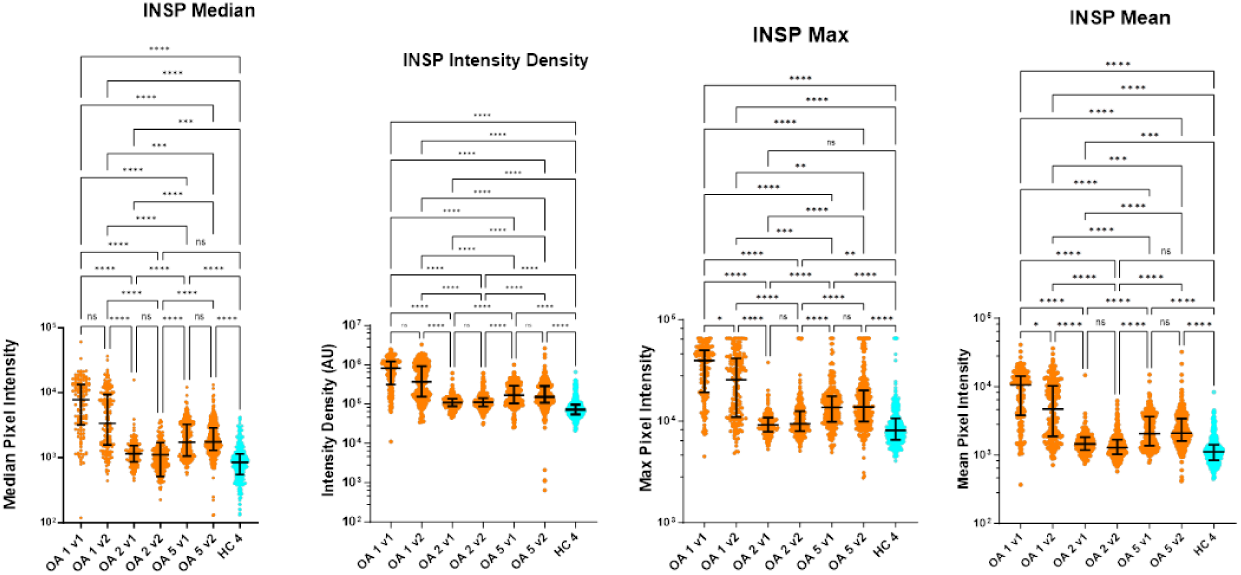
IgG purified from visit 1 (baseline) and visit 2 (post-exercise) serum of OA patients (OA 1 = Pt1, OA 2 = Pt2, OA 5 = Pt5, and a healthy control tested for SGC binding.

**Figure 4.**
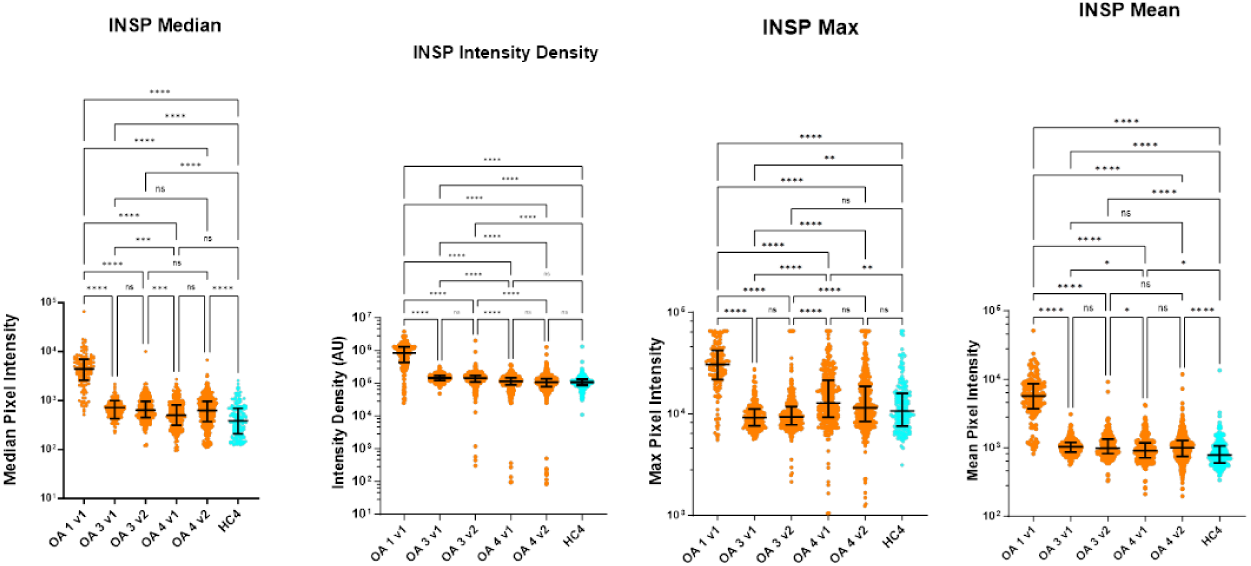
IgG purified from visit 1 (baseline) and visit 2 (post-exercise) serum of OA patients (OA 3 = Pt3, OA 4 = Pt4, and a healthy control tested for SGC binding. OA1 visit 1 serum served as a positive control.

## Discussion

In this exploratory study, we used complementary circulating protein, whole-blood transcriptomic and SGC-binding IgG measurements to investigate biological features associated with heterogeneity in pain response to lifestyle interventions in knee OA. No individual proteomic or transcriptomic association remained significant after correction for multiple testing but several nominal associations were identified, including baseline IL-10RA, longitudinal IL-17A and MMP9 expression. The findings should therefore be considered hypothesis-generating rather than evidence of established molecular markers of treatment response. However, the observed inflammatory and extracellular-matrix pathways may warrant further investigation.

The most consistent signal across the proteomic analyses was the direction of association between longitudinal changes in IL-17A and pain improvement. In INSPIRE, greater increases in circulating IL-17A were nominally associated with greater improvement in pain, and a similar direction of association was observed in the independent WebEx cohort. Neither association remained significant following FDR correction, and the consistency between cohorts should therefore be interpreted cautiously rather than as evidence of replication. Nevertheless, IL-17A is of biological interest because IL-17 signalling has been implicated in synovial inflammation, cartilage degradation and inflammatory mediator production in OA [14]. Studies in knee OA have reported higher serum and synovial IL-17A concentrations in association with pain and functional impairment [15], although findings across studies are inconsistent. A recent systematic review similarly concluded that circulating IL-17 shows suggestive but inconsistent associations with OA-related pain, whereas evidence for synovial IL-17 is more consistent [16].

The direction of the longitudinal association observed in the present study is therefore not straightforward to interpret. Greater increases in IL-17A were associated with greater pain improvement, despite IL-17A generally being considered a pro-inflammatory mediator associated with OA pathology. This suggests that changes in circulating IL-17A during lifestyle intervention may not directly reflect changes in joint inflammation. Rather, they could represent broader immune adaptation accompanying clinical improvement. This interpretation, although speculative given that the association did not survive multiple-testing correction, is consistent with evidence that exercise interventions can produce heterogeneous changes in circulating inflammatory markers in OA. For example, exercise has been associated with reductions in IL-6 in some studies, particularly where exercise is combined with weight loss [17, 18], while other trials have observed improvements in pain without corresponding changes in circulating pro-inflammatory cytokines [19]. More broadly, mediation analyses from the ADAPT trial suggested that changes in systemic inflammatory markers accounted for part of the effect of diet and exercise on pain, supporting a potential role for systemic inflammation in treatment response [20].

Baseline IL-10RA was nominally associated with non-response under both pain-response definitions in INSPIRE, although neither association survived correction for multiple testing. IL-10 signalling is generally considered anti-inflammatory and has been implicated in protection against OA-associated inflammation and pain, although evidence specifically linking circulating IL-10RA to clinical pain response is limited. Interestingly, IL10RA has also been identified among differentially expressed genes in OA synovial tissue and in recent genetic analyses of inflammatory proteins and OA, but its precise role appears context dependent [21]. The present finding should therefore not be interpreted as evidence that IL-10RA directly influences treatment response. Instead, it may reflect differences in immune regulation between individuals, a possibility that would require investigation in larger and independently powered studies.

OSM and TNFSF14 were associated nominally with non-response in WebEx but did not reproduce the INSPIRE findings and did not remain significant following multiple-testing correction. OSM is of particular biological interest because it has been linked to an inflammatory OA phenotype, synovitis and extracellular-matrix remodelling, although it appears to be present only in a subset of patients [22]. The lack of replication of the specific baseline proteins between cohorts therefore may be consistent with the recognised molecular heterogeneity of OA, rather than necessarily indicating that individual inflammatory proteins are irrelevant to pain response. However, given the exploratory nature of the analyses and the absence of FDR-significant associations, this interpretation cannot be distinguished from chance variation between cohorts.

MMP9 was the strongest transcriptomic signal under both definitions of pain response, showing a greater longitudinal increase in the responder group. Although neither association survived multiple-testing correction, the consistency across the two responder thresholds makes MMP9 a potentially interesting candidate for further study.

MMP9 has an established role in OA biology, particularly in extracellular-matrix degradation and inflammatory tissue remodelling [23]. Increased MMP9 has been reported in OA serum and synovial fluid, and circulating MMP9 expression has also been associated with clinical symptom severity [24]. MMP9-mediated extracellular-matrix remodelling may therefore provide a mechanistic link between joint tissue biology and pain-related phenotypes. However, our finding concerns a change in whole-blood MMP9 transcription rather than MMP9 activity within the joint. The direction of association with treatment response was also not what would be predicted simply from considering MMP9 as a marker of OA tissue degradation. This again suggests that systemic transcriptional changes may reflect broader biological responses to intervention rather than directly representing pathogenic activity within the joint. Alternatively, the nominal association may represent a chance finding arising from the multiple comparisons performed.

The exploratory SGC-binding experiments addressed a different possibility: that an immune-mediated mechanism involving peripheral sensory structures could identify individuals with particularly poor or paradoxical responses to exercise. This hypothesis was motivated by work demonstrating that IgG from people with fibromyalgia can bind satellite glial cells and sensory neurons and, following transfer to mice, induce sensory hypersensitivity [10].

One participant with marked worsening of pain showed substantially increased baseline SGC-binding IgG, providing an intriguing individual-level observation. However, this pattern was not observed in the other selected poor responders, including a participant with marked pain worsening but low baseline binding. Furthermore, paired samples did not show a consistent change in SGC-binding after intervention. These findings therefore do not support a simple model in which circulating SGC-binding IgG is a general predictor of exercise-related pain worsening or improvement in knee OA. Given the very small, deliberately selected sample, however, they cannot exclude a role for IgG antibodies in a subset of patients. Of note, a role for the immunoglobulin IgM isotype [25] has been demonstrated in a mouse OA model. Larger studies with systematic sampling, human sensory-tissue validation and functional assays would be required to determine whether SGC-reactive or other antibodies define a distinct OA pain phenotype.

## Conclusions

Taken together, this exploratory multi-modal analysis did not identify a robust molecular signature of variation in pain response to lifestyle intervention in knee OA. No individual proteomic or transcriptomic association survived correction for multiple testing, and the exploratory SGC-binding experiments provided no evidence of a consistent autoantibody phenotype associated with poor clinical response.

Several nominal findings, including longitudinal IL-17A associations in both cohorts and MMP9 expression under both pain-response definitions, provide potential leads for future investigation involving interacting biological processes rather than a single circulating biomarker. This is consistent with current models of OA pain, which recognise contributions from synovial inflammation, tissue remodelling and peripheral and central nociceptive mechanisms, with substantial discordance between structural disease and pain severity. However, the absence of FDR-significant associations is important. The present sample sizes, particularly for the transcriptomic and autoantibody analyses, limit power, and the Olink and whole-blood measurements provide only indirect measures of processes occurring within the joint and peripheral nervous system.

A strength of the study was the use of complementary proteomic, transcriptomic and autoantibody approaches to examine several potential biological explanations for inter-individual variation in treatment response. Future studies should test the observed candidate signals prospectively in larger, independently powered cohorts, ideally integrating circulating biomarkers with synovial or imaging measures, sensory phenotyping and longitudinal clinical outcomes. Such studies will be important for determining whether the nominal associations observed for inflammatory and extracellular-matrix pathways reflect reproducible biological differences between pain-response phenotypes in people with knee OA or chance findings arising from exploratory analyses.

## Data Availability

All data produced in the present study are available upon reasonable request to the authors

